# Automated Transformation of Unstructured Cardiovascular Diagnostic Reports into Structured Datasets Using Sequentially Deployed Large Language Models

**DOI:** 10.1101/2024.10.08.24315035

**Authors:** Sumukh Vasisht Shankar, Lovedeep S Dhingra, Arya Aminorroaya, Philip Adejumo, Girish N Nadkarni, Hua Xu, Cynthia Brandt, Evangelos K Oikonomou, Aline F Pedroso, Rohan Khera

## Abstract

**Background:** Rich data in cardiovascular diagnostic testing are often sequestered in unstructured reports, with the necessity of manual abstraction limiting their use in real-time applications in patient care and research.

**Methods:** We developed a two-step process that sequentially deploys generative and interpretative large language models (LLMs; Llama2 70b and Llama2 13b). Using a Llama2 70b model, we generated varying formats of transthoracic echocardiogram (TTE) reports from 3,000 real-world echo reports with paired structured elements, leveraging temporal changes in reporting formats to define the variations. Subsequently, we fine-tuned Llama2 13b using sequentially larger batches of generated echo reports as inputs, to extract data from free-text narratives across 18 clinically relevant echocardiographic fields. This was set up as a prompt-based supervised training task. We evaluated the fine-tuned Llama2 13b model, HeartDx-LM, on several distinct echocardiographic datasets: (i) reports across the different time periods and formats at Yale New Haven Health System (YNHHS), (ii) the Medical Information Mart for Intensive Care (MIMIC) III dataset, and (iii) the MIMIC IV dataset. We used the accuracy of extracted fields and Cohen’s Kappa as the metrics and have publicly released the HeartDX-LM model.

**Results:** The HeartDX-LM model was trained on randomly selected 2,000 synthetic echo reports with varying formats and paired structured labels, with a wide range of clinical findings. We identified a lower threshold of 500 annotated reports required for fine-tuning Llama2 13b to achieve stable and consistent performance. At YNHHS, the HeartDx-LM model accurately extracted 69,144 out of 70,032 values (98.7%) across 18 clinical fields from unstructured reports in the test set from contemporary records where paired structured data were also available. In older echo reports where only unstructured reports were available, the model achieved 87.1% accuracy against expert annotations for the same 18 fields for a random sample of 100 reports. Similarly, in expert-annotated external validation sets from MIMIC-IV and MIMIC-III, HeartDx-LM correctly extracted 201 out of 220 available values (91.3%) and 615 out of 707 available values (87.9%), respectively, from 100 randomly chosen and expert annotated echo reports from each set.

**Conclusion:** We developed a novel method using paired large and moderate-sized LLMs to automate the extraction of unstructured echocardiographic reports into tabular datasets. Our approach represents a scalable strategy that transforms unstructured reports into computable elements that can be leveraged to improve cardiovascular care quality and enable research.

## INTRODUCTION

Electronic health records (EHR) offer invaluable insights into optimizing cardiovascular care and driving healthcare research.^1–3^ In the EHR, data streams that are most amenable to scalable applications include those available as structured tabular data. Therefore, despite their critical role in defining disease conditions, diagnostic testing such as imaging is often available only as unstructured free-text narratives and remains underutilized in disease phenotyping.^4^ This gap underscores the pressing need for novel strategies to transform unstructured data into structured data elements, thus enhancing the impact and scalability of health applications that can leverage these rich data.

Prior work to transform unstructured into structured data has primarily focused on extracting isolated data elements,^5–9^ with the need to develop pipelines to extract as more data streams are needed. The emergence of large language models (LLMs) as foundation models for language processing has demonstrated impressive properties for parsing text with limited domain-specific development but are limited by the high computational requirements associated with their deployment.^10^ On the other hand, the scarcity of annotated unstructured-structured data limits the development of computationally efficient models. Consequently, there is a critical unmet need for novel approaches capable of efficiently transforming clinical notes into tabular data.

To address this, we propose a domain-specific and computationally efficient approach leveraging sequentially deployed LLMs, where we use a larger open-source model to generate synthetic training examples for fine-tuning a smaller model, which enables the development of a generalizable tool for converting imaging reports to tabular data. We use reports of transthoracic echocardiograms (TTEs) as the use case for this application.

## METHODS

The study was reviewed by the Yale Institutional Review Board, which waived the need for informed consent, as it represents a secondary analysis of existing data.

### Study Overview

We developed and fine-tuned a lightweight language model, HeartDx-LM, to extract clinically relevant diagnostic information from unstructured TTE reports. We trained the model using different text structures leveraging temporal variations in the free-text narratives of the reports to introduce this variation. The process involved generating synthetic reports using reports in a single format where all information was also available as tabular data. These reports were regenerated into different formats using examples from those formats as prompts to a Llama2 70-billion-parameter model. These synthetically adapted echo reports were then used to fine-tune a moderate-sized Llama2 13-billion-parameter model to extract a comprehensive set of quantitative, semi-quantitative, and qualitative diagnostic information from unstructured clinical reports (**Figure 1**).

**Figure 1:**
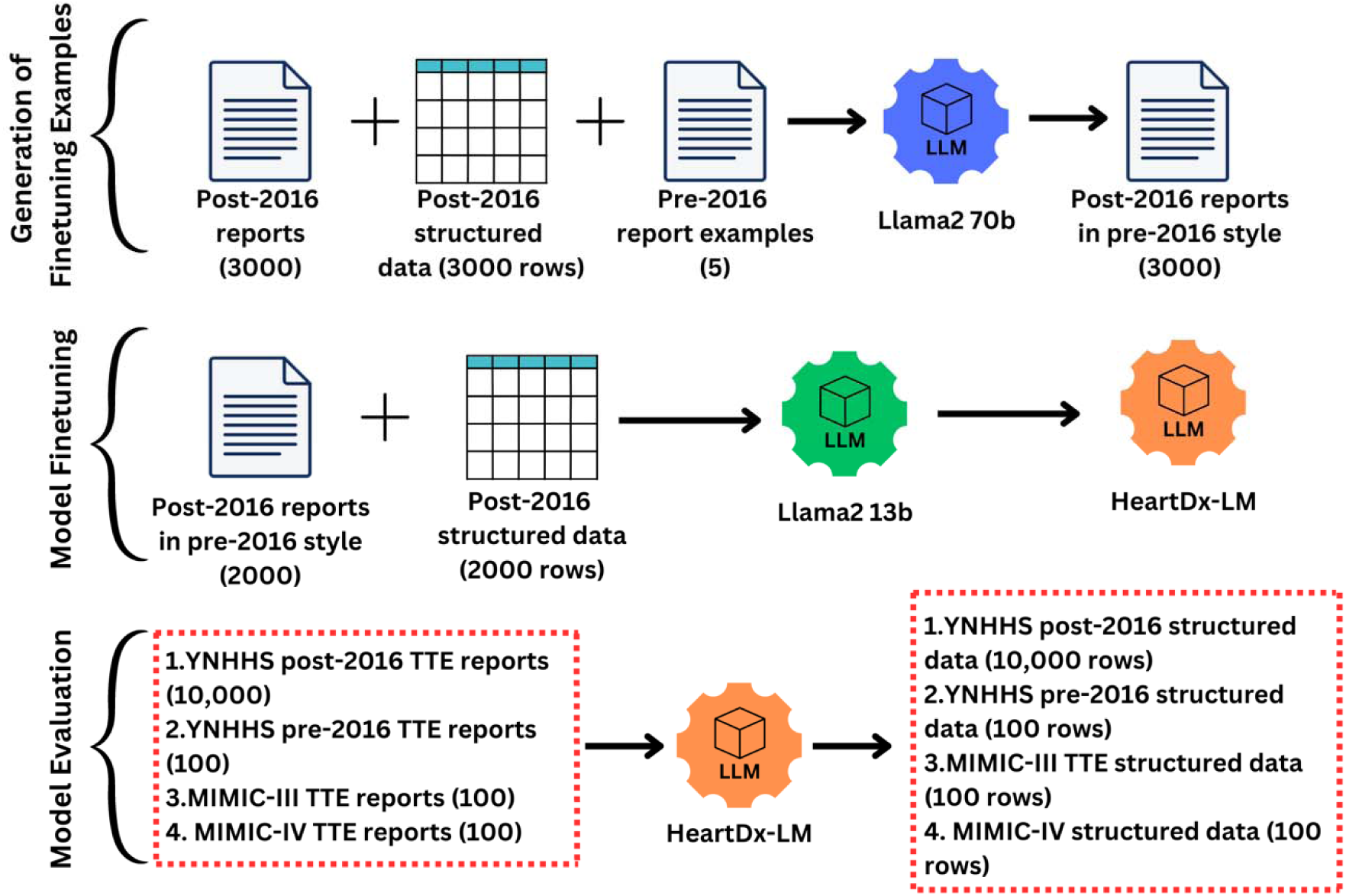
Model Development Approach and Study Design. **Abbreviations:** YNHHS, Yale New Haven Health System; MIMIC, Medical Information Mart for Intensive Care

### Data Sources

We used data from the Yale New Haven Health System (YNHHS) EHR, a large academic health system catering to a diverse population in New Haven County, one of the most representative counties in the US. Since 2016, the free-text imaging reports for TTEs have been linked with structured tabular values for the cardiologist-defined echocardiographic features. The linked structured dataset consisted of clinical and operational labels, of which we selected 18 based on their broad coverage of key conditions. These included ejection fraction (EF), global longitudinal strain, interventricular septal thickness (IVSd), aortic valve (AV) and mitral valve (MV) structure, and qualitative or quantitative features associated with AV and MV stenosis/regurgitation, including left ventricular outflow tract (LVOT) peak velocity and peak gradient, AV peak velocity and mean gradient, AV area by continuity, and AV area index. A brief overview of the data fields is included in **Table 1**. This dataset of 10,000 reports paired with corresponding structured labels was used for model evaluation (test set).

**Table 1:**
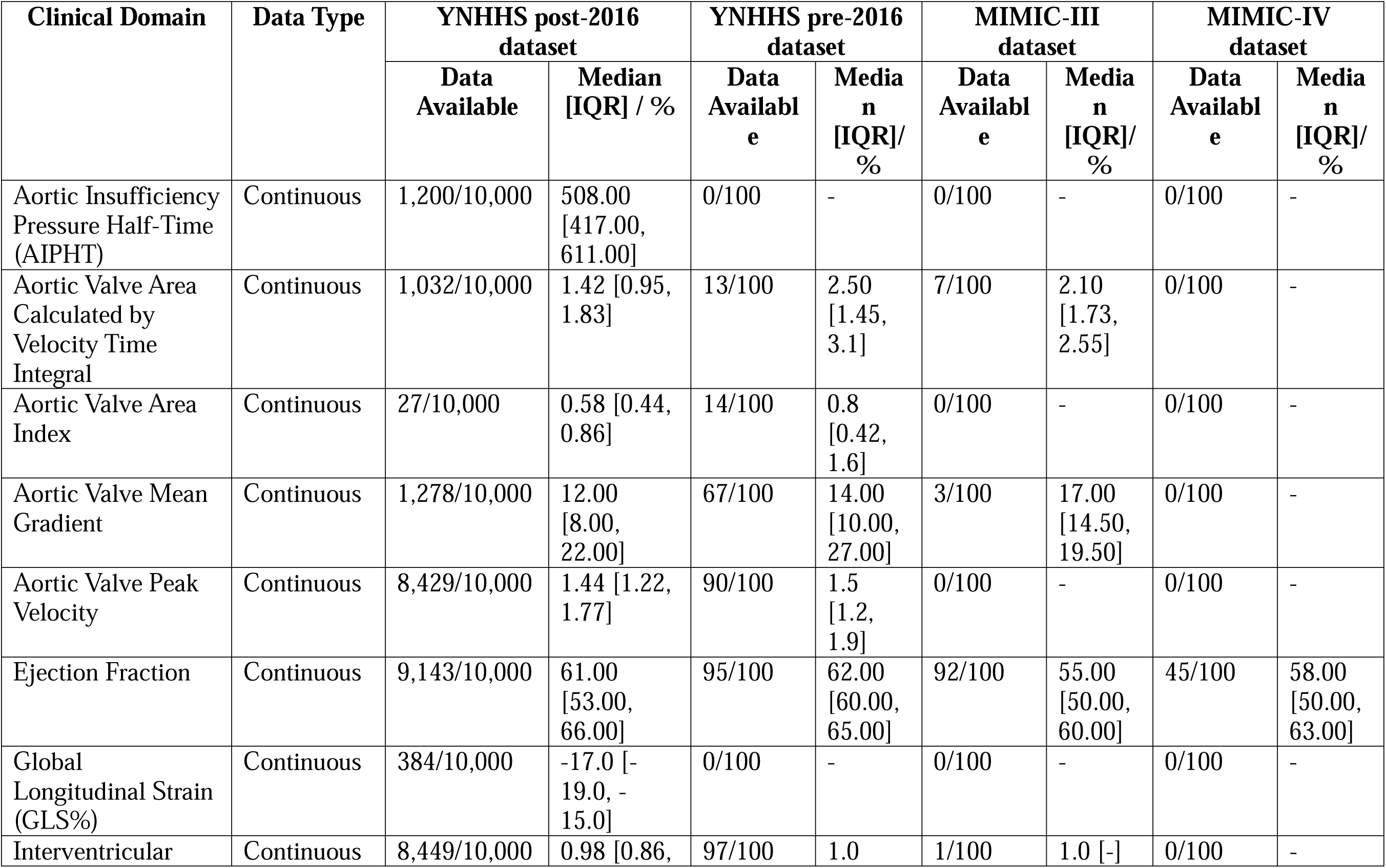
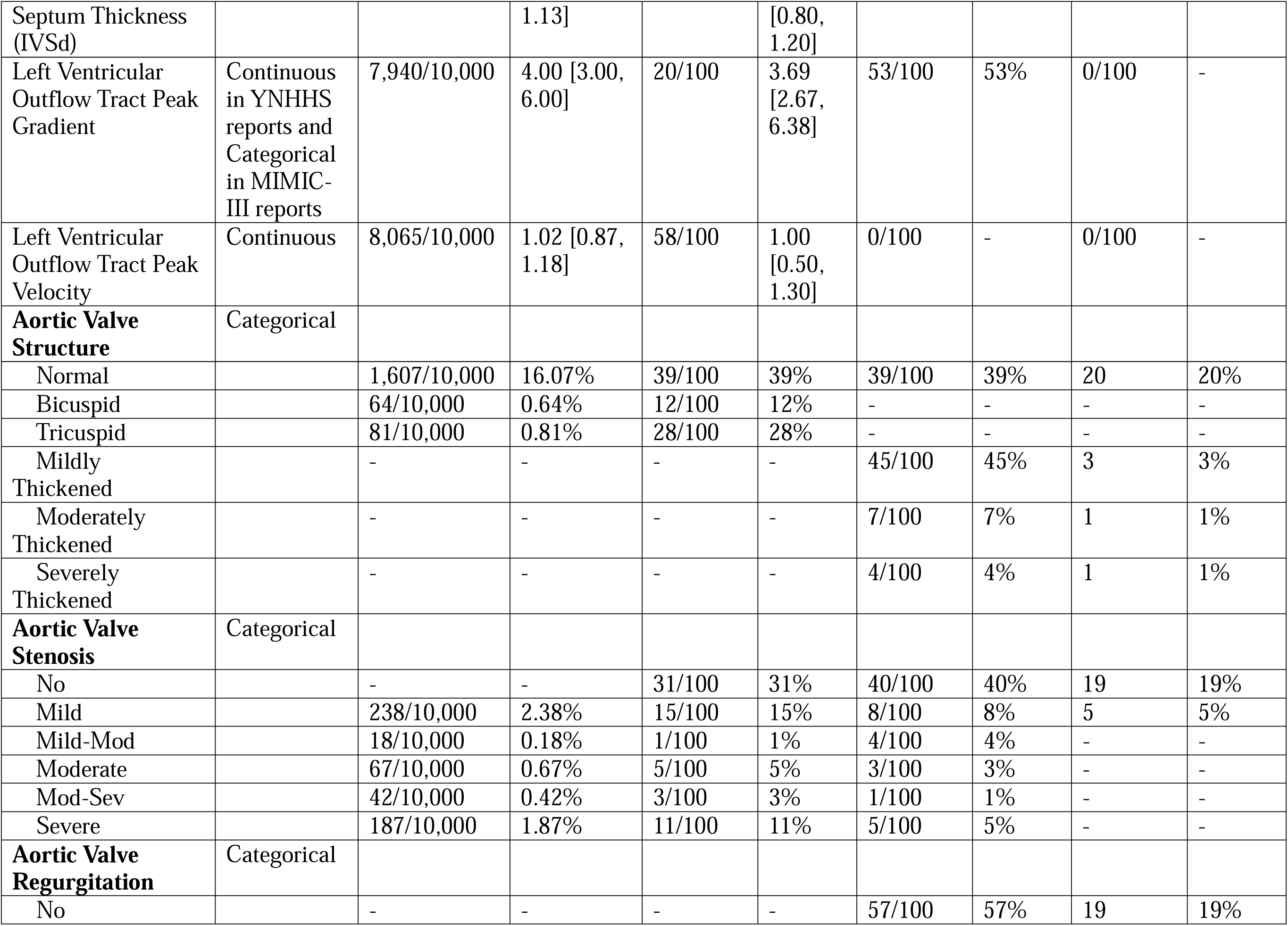
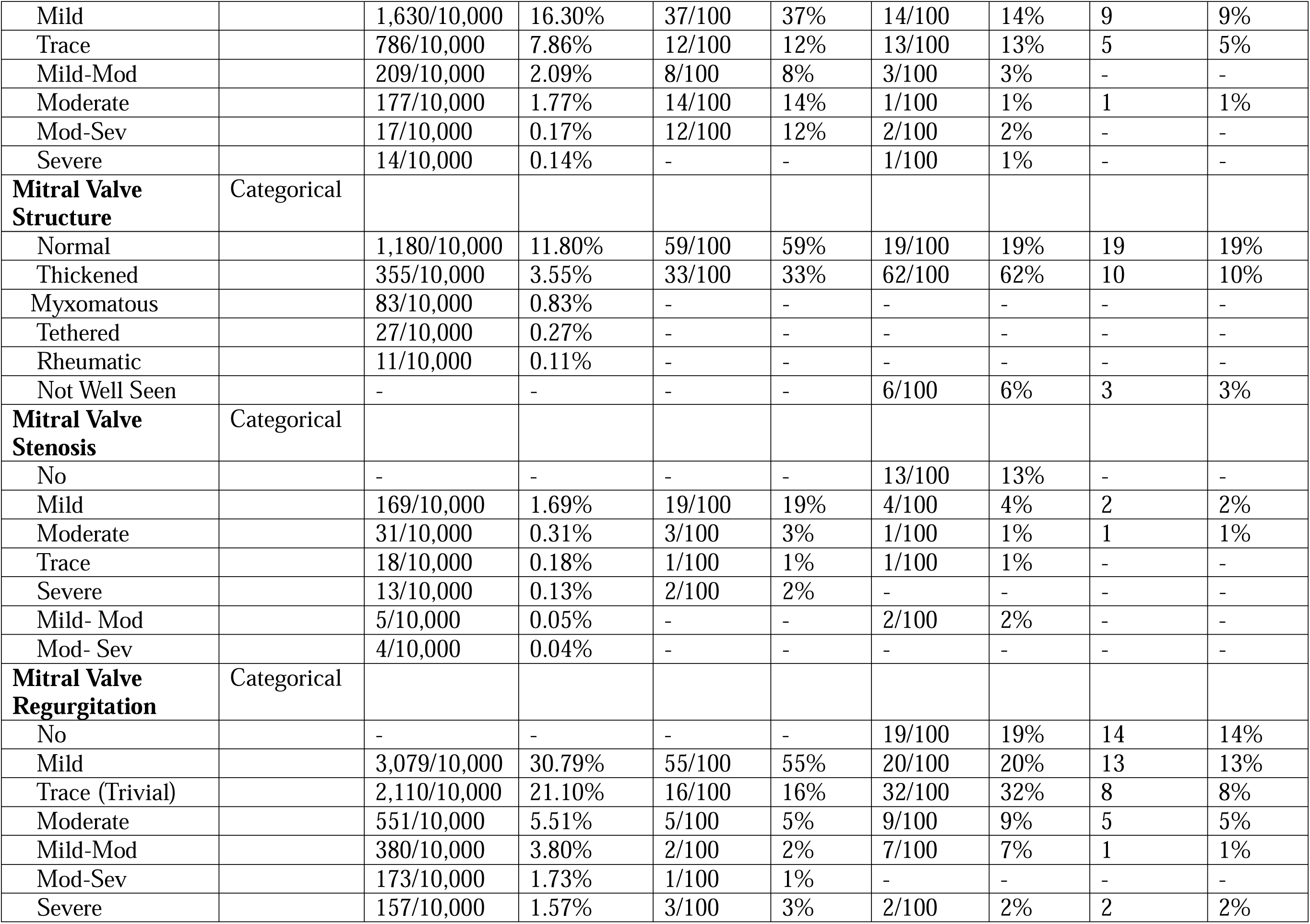
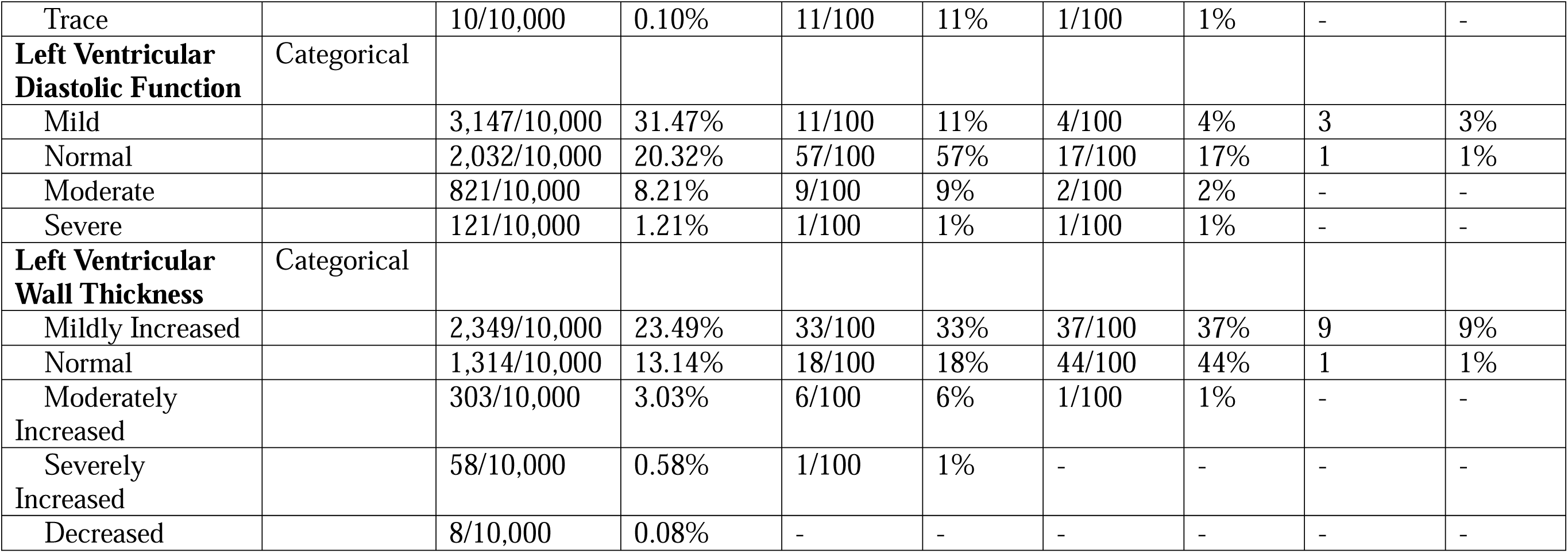
Data Summarization of the train, test, and validation datasets.

Structurally distinct TTE reports from MIMIC-III and MIMIC-IV datasets were used for external validation of our digitization approach.^11,12^ MIMIC-III comprises deidentified EHR data from over forty thousand patients with a hospitalization that included an intensive care unit stay at the Beth Israel Deaconess Medical Center between 2001 and 2012. The data represents broad EHR fields spanning demographics, laboratory test results, procedures, medications, caregiver notes, imaging reports, and mortality. MIMIC-IV is an updated version of the MIMIC-III database, which incorporates data up to 2019 and includes hospitalizations with emergency department visits. The current study leveraged echocardiographic reports from both MIMIC-III and MIMIC-IV. Representative examples of various report types from the different sources are included in **Supplemental Table S1**.

### Model Development: Overall approach

We designed a two-step approach to convert unstructured TTE reports to structured data (digitize) using LLMs. All TTE reports in the YNHHS dataset post-2016 had corresponding clinician-annotated tabular data, which provided us with a training set without the need for manual annotation. However, reports from before this time (pre-2016) were only available in a free-text format without corresponding tabular data. Moreover, the data had several variations in free text reporting across echocardiographers. From a randomly selected 3,000 pre-2016 YNHHS reports, we observed 5 different reporting formats (**Supplemental Table 2**). We randomly chose one report from each unique reporting format to encode this variation in our training set.

The utilization of Llama for finetuning, as opposed to alternative LLMs, was driven by considerations of its parameter efficiency, domain-specific architecture, and applicability to the medical text processing domain. By prioritizing factors such as model performance, accessibility, and computational power required to finetune the model, we aimed to optimize the efficiency and effectiveness of the finetuning process. This also included the ability to quantize Llama models into a 4-bit configuration for reduction in model size and memory usage.^13–15^

### First-stage development: Finetuning Llama2 70b

In the initial phase, we fine-tuned the Llama2 70-billion-parameter LLM to generate TTE reports from the structured data in the post-2016 reports with syntactical characteristics – including the five formatting variations – of the pre-2016 reports. We trained the model on a dataset of 3,000 paired examples from the post-2016 dataset to create unstructured data that faithfully represents the format of pre-2016 reports for subsequent fine-tuning and testing (Prompt template – **Supplemental Table 2**).

The task of restructuring reports from post-2016 format to pre-2016 format involved fine-tuning the pre-trained Llama2-70b model to recognize and replicate the syntactical and structural elements of various pre-2016 reporting formats. The model was trained on a curated dataset containing examples of both post-2016 and pre-2016 reports. The dataset was carefully prepared, ensuring the pre-2016 reports represented multiple versions and styles to comprehensively expose the model to various data formats. The model was fine-tuned over two epochs with a batch size of 4, using the Adam optimizer with a learning rate of 10^-5^. The choice of hyperparameters, including the learning rate and epochs, was based on commonly adopted practices for fine-tuning large language models.^16,17^ The fine-tuning process was monitored in a validation set, with early stopping of fine-tuning when validation loss did not improve for 5 consecutive evaluation steps to prevent overfitting. This approach was implemented to ensure that the model generalizes well to unseen data while maintaining high accuracy on the training set.

### Second-stage development: Finetuning Llama2 13b

After the initial step, we used the restructured reports created with the Llama2 70-billion-parameter LLM to train a Llama2 13-billion-parameter LLM. We trained the model on a subset of 2,000 regenerated TTE reports, each paired with clinician-annotated tabular data. This approach enabled the model to learn from TTE reports that vary in formats while still being able to use corresponding clinician-annotated tabular data as the gold standard. This led to our model, HeartDx-LM, which is tailored to extracting structured fields from free-text narratives of TTE reports across the selected 18 clinical variables without requiring the large computational infrastructure needed for the 70-billion-parameter model. HeartDx-LM was trained to discern and extract critical information (Prompt table – **Supplemental Table 4**). We have made the model publicly available on HuggingFace at https://huggingface.co/CarDSLab/HeartDX-LM.

We also digitized the TTE reports using a non-finetuned Llama2-13b model (zero-shot Llama) to compare its performance with its finetuned counterparts.

### Evaluation

We conducted a comprehensive evaluation of the model’s performance across four distinct datasets – post-2016 YHNNS TTE reports (internal held-out test set), pre-2016 YNHHS reports, MIMIC-III TTE reports, and MIMIC-IV TTE reports.

Firstly, we employed a held-out set comprising 10,000 post-2016 YNHHS reports sourced from the YNHHS EHR. These reports were accompanied by their corresponding structured fields, allowing for direct comparison and assessment of the model’s proficiency in extracting structured data from contemporary clinical narratives.

In addition to the post-2016 YNHHS dataset, we also examined the model’s performance on 100 pre-2016 YNHHS reports and 100 reports each from the MIMIC-III and MIMIC-IV datasets. The pre-2016 YNHHS dataset used for model evaluation were a distinct set from the one used to develop synthetic examples and had clinical labels manually extracted by three clinical experts. The TTE reports from MIMIC-III were obtained from the EchoNotes Structured Database, which also includes echocardiogram reports from the intensive care unit.^18,19^ In the MIMIC-IV dataset, reports were retrieved from discharge summaries that contained TTE report summaries. This structured echocardiogram database included key measures of cardiac structure and function, such as ejection fraction (EF), aortic valve (AV) and mitral valve (MV) structure, and qualitative or quantitative features associated with AV and MV stenosis/regurgitation. The other structured fields of interest like interventricular septal thickness (IVSd), left ventricular outflow tract (LVOT) peak velocity and peak gradient, AV peak velocity and mean gradient, AV area by continuity, and AV area index were derived through manual annotation by three clinical experts, who collaboratively established an annotation scheme delineating the criteria for extracting values for each of the 18 clinical variables. Each report was evaluated based on its constituent sentences, and the clinicians’ annotations were aggregated to create a gold standard for evaluating the model’s performance. This ensured the accuracy and reliability of the ground truth.

In addition to evaluating the performance of our fine-tuned models, we also employed the Llama2-13b model without fine-tuning as a comparator to assess its capability in extracting structured data from clinical narratives without prior training on our datasets. This allowed us to benchmark the performance of our approach against the out-of-box (or zero-shot) performance of the Llama2-13b model.

### Determining Optimal Training Data Volume for Model Fine-tuning

We investigated the impact of training data volume on model performance by fine-tuning multiple iterations of the Llama2-13b model with progressively increasing numbers of training reports (100, 200, 500, 1,000, and 2,000). We evaluated the model’s performance on the held-out post-2016 YNHHS dataset for each iteration. This comprehensive evaluation framework aimed to elucidate the model’s capabilities and provide insights into the optimal training data volume required for achieving robust performance in extracting structured information from diverse clinical narratives. We used a statistical benchmark of 95% accuracy to define robust performance.

### Statistical Analysis

We assessed HeartDx-LM’s performance using the accuracy of extracted values for both continuous and categorical variables. We reported the overall extraction accuracies and accuracies for individual clinical variables compared against the ground truth annotations. Extraction accuracy was defined as the percentage of values correctly extracted by the model, with incorrect and failed extractions rates also reported. Additionally, Cohen’s kappa statistic was used to evaluate the agreement between the model’s extractions and the ground truth for both categorical and continuous variables. Specifically, continuous variables were categorized into discrete classes for the Kappa analysis. Each continuous variable in the training dataset was labeled as either 1 for available values or 0 for missing values. For the digitized dataset, the continuous variables were labeled based on their comparison with the original dataset: a label of 1 for values that were available in both the training and digitized datasets and correctly extracted, a label of 0 for values that were missing in both datasets (correctly identified as missing), a label of 2 for values available in the training dataset but extracted incorrectly in the digitized dataset, and a label of 3 for values available in the training dataset but missing in the digitized dataset. We calculated Cohen’s Kappa for each continuous variable independently, measuring the agreement between the original and digitized labels across these four categories. The Kappa statistic was computed using the formula *K* = (*P_o_* –P*_e_*)/(1 – P*_e_*), where P*_O_* is the observed agreement between the two datasets and P*_e_* is the expected agreement by chance. For multiclass Kappa, the observed and expected agreements considered all four categories to provide a comprehensive measure of agreement. By categorizing the continuous variables and then applying Cohen’s Kappa, we ensured that the agreement between the original and digitized datasets was evaluated robustly, accounting for both correct and incorrect extractions as well as missing data.

The Cohen’s Kappa statistic metric accounts for the possibility of agreement occurring by chance, providing a more robust measure of the model’s reliability.^20^ A kappa value closer to 1 indicates a high level of agreement, while a value closer to 0 suggests agreement is no better than chance.

## RESULTS

### Study Population

There were 8,612 unique patients with 10,000 post-2016 YNHHS reports in the test set, with a median age of 73.0 (IQR, 62.0 – 85.0) years, including 5,013 (50.1%) women, 694 (6.9%) non-Hispanic Blacks, 88 (0.9%) Hispanics, and 52 (0.5%) of Asian race. The range of distribution of clinical features (across both development and validation cohorts) are provided in **Table 1**.

### Zero-shot model performance

The zero-shot Llama2-13b model generated fragmented, inconsistent, or irrelevant responses, resulting in incomplete and inaccurate extractions. This resulted in 0% extraction accuracy across all the 18 clinical variables. An example of zero-shot model prompt and response is shown in **Supplemental Table 5**.

### Model performance in the held-out test set (post-2016 YNHHS dataset)

The HeartDx-LM model extracted 69,144 out of 70,032 values, yielding an accuracy rate of 98.7% and Cohen’s Kappa value of 0.99. The accuracy rate was consistent across both continuous (45712/46387 – 98.5%) and categorical (23432/23645 – 99.1%) variables. The model incorrectly extracted 480 (0.7%) values and did not extract 408 (0.6%) values. The inaccurate values were most frequent for EF (97/9143), followed by AV peak velocity (96/8429), LVOT peak velocity (74/8065), and IVSd (48/8449).

Across continuous variables, the accuracy of the model for key clinical variables like EF, LVOT peak velocity, and AV peak velocity were 97.3% (8,902/9,143), 98.8% (7,969/8,065), and 98.8% (8,331/8,429), respectively. For key categorical variables of AV structure, AV regurgitation, MV regurgitation, and LV wall thickness, the accuracies were 98.6% (1,727/1,752), 99.1% (2,813/2,833), 99.7% (6,440/6,460), and 97.7% (3,938/4,032), respectively (**Table 2**).

**Table 2:** Model Performance Evaluation of HeartDX-LM on the held-out test set.

| **Metrics** | **Available Values** | **Correct: N (%)**<br>**(Extracted value matches original value)** | **Incorrect: N (%)**<br>**(Extracted value differs from original value)** | **Failed: N (%)**<br>**(Value not extracted)** | **Cohen's Kappa** |
| --- | --- | --- | --- | --- | --- |
| **Column** |  |  |  |  |  |
| **All variables** | 70,032 | 69,144 (98.7) | 480 (0.7) | 408 (0.6) | 0.99 |
| **AIPHT** | 1,200 | 1,173 (98) | 0 (0) | 27 (2) | 0.98 |
| **AVA Cont VTI** | 1,032 | 981 (95.3) | 0 (0) | 51 (4.7) | 0.97 |
| **AVA Index** | 27 | 27 (100) | 0 (0) | 0 (0) | 1.0 |
| **AV Mn Grad** | 1,728 | 1,680 (97.2) | 25 (1.4) | 23 (1.4) | 0.98 |
| **AV Pk Vel** | 8,429 | 8,331 (98.8) | 96 (1.1) | 2 (0.1) | 0.96 |
| **Ejection Fraction** | 9,143 | 8,902 (97.3) | 97 (1.1) | 144 (1.6) | 0.87 |
| **GLS%** | 384 | 383 (99.7) | 0 (0) | 1 (0.3) | 0.99 |
| **IVSd** | 8,449 | 8,401 (99.4) | 48 (0.6) | 0 (0) | 0.98 |
| **LVOT Pk Grad** | 7,940 | 7,868 (99.1) | 43 (0.6) | 29 (0.3) | 0.98 |
| **LVOT Pk Vel** | 8,065 | 7,969 (98.8) | 74 (0.9) | 22 (0.3) | 0.97 |
| **AV Structure** | 1,752 | 1,727 (98.6) | 25 (1.7) | 0 (0) | 0.99 |
| **AV Stenosis** | 552 | 552 (100) | 0 (0) | 0 (0) | 1.0 |
| **AV Regurgitation** | 2,833 | 2,813 (99.1) | 0 (0) | 20 (0.9) | 0.99 |
| **MV Structure** | 1,656 | 1,656 (100) | 0 (0) | 0 (0) | 1.0 |
| **MV Stenosis** | 240 | 240 (100) | 0 (0) | 0 (0) | 1.0 |
| **MV Regurgitation** | 6,460 | 6,440 (99.7) | 0 (0) | 20 (0.3) | 0.99 |
| **LV Diastolic Function** | 6,121 | 6,074 (99.2) | 47 (0.8) | 0 (0) | 0.99 |
| **LV Wall Thickness** | 4,032 | 3,938 (97.7) | 25 (0.6) | 69 (1.7) | 0.98 |

Abbreviations: AVA Cont VTI, Aortic Valve Area Calculated by Velocity Time Integral ; AVA Index, Aortic Valve Area Index; AV Mn Grad, Aortic Valve Mean Gradient; AIPHT, Aortic Insufficiency Pressure Half-Time; AV Pk Vel, Aortic Valve Peak Velocity; AV Regurgitation, Aortic Valve Regurgitation; AV Stenosis, Aortic Valve Stenosis; AV Structure, Aortic Valve Structure; GLS, Global Longitudinal Strain; IVSd, Interventricular Septum Thickness; ; LV Diastolic Function, Left Ventricular Diastolic Function; LVOT Pk Grad, Left Ventricular Outflow Tract Peak Gradient; LVOT Pk Vel, Left Ventricular Outflow Tract Peak Velocity; LV Wall Thickness, Left Ventricular Wall Thickness; MV Regurgitation, Mitral Valve Regurgitation; MV Stenosis, Mitral Valve Stenosis; MV Structure, Mitral Valve Structure.

### Model performance in pre-2016 reports

In the 100 randomly sampled and expert-annotated pre-2016 reports, HeartDx-LM achieved an overall accuracy rate of 87.1% (extracting 909 out of the 1044 values), and Cohen’s Kappa value of 0.86 across 18 clinical variables when compared against manually annotated labels. The model incorrectly extracted 11 (1.1%) values and failed to extract 124 (11.9%) values. The inaccurate values were most frequent for AV mean gradient (3/67), followed by MV structure (3/92), MV stenosis (2/25), MV regurgitation (1/93) and LV diastolic function (1/78).

HeartDx-LM maintained a high accuracy across both continuous (407/454 – 89.6%) and categorical (502/590 – 85.1%) variables. Accuracy of the model across key continuous variables, of EF, LVOT peak velocity, and AV peak velocity were 90.5% (86/95), 86.2% (50/58), and 92.2% (83/90), respectively. For key categorical variables of AV structure, AV regurgitation, MV regurgitation, and LV wall thickness, the accuracies were 83.2% (79/95), 95.2% (79/83), 93.5% (87/93), and 56.9% (33/58), respectively (**Table 3**).

**Table 3:** Model Performance Evaluation of HeartDX-LM on Pre-2016 Reports.

| Metrics | Available Values | Correct: N (%)<br>(Extracted value matches original value) | Incorrect: N (%)<br>(Extracted value differs from original value) | Failed: N (%)<br>(Value not extracted) | Cohen's Kappa |
| --- | --- | --- | --- | --- | --- |
| Column |  |  |  |  |  |
| **All variables** | 1044 | 909 (87.1) | 11 (1.1) | 124 (11.9) | 0.86 |
| **AIPHT** | 0 | - | - | - | - |
| **AVA Cont VTI** | 13 | 10 (76.9) | 0 (0) | 3 (23.1) | 0.87 |
| **AVA Index** | 14 | 12 (85.7) | 0 (0) | 2 (14.3) | 0.92 |
| **AV Mn Grad** | 67 | 59 (88) | 3 (4.4) | 5 (74.6) | 0.84 |
| **AV Pk Vel** | 90 | 83 (92.2) | 1 (1.1) | 6 (6.6) | 0.71 |
| **Ejection Fraction** | 95 | 86 (90.5) | 0 (0) | 9 (9.5) | 0.50 |
| **GLS%** | 0 | - | - | - | - |
| **IVSd** | 97 | 90 (92.8) | 0 (0) | 7 (7.2) | 0.44 |
| **LVOT Pk Grad** | 20 | 17 (85) | 0 (0) | 3 (15) | 0.91 |
| **LVOT Pk Vel** | 58 | 50 (86.2) | 0 (0) | 8 (13.8) | 0.85 |
| **AV Structure** | 95 | 79 (83.2) | 0 (0) | 16 (16.8) | 0.35 |
| **AV Stenosis** | 66 | 56 (84.8) | 0 (0) | 10 (15.2) | 0.81 |
| **AV Regurgitation** | 83 | 79 (95.2) | 0 (0) | 4 (4.8) | 0.87 |
| **MV Structure** | 92 | 80 (86.9) | 3 (3.2) | 9 (9.8) | 0.53 |
| **MV Stenosis** | 25 | 22 (88) | 2 (8) | 1 (4) | 0.92 |
| **MV Regurgitation** | 93 | 87 (93.5) | 1 (1.1) | 5 (5.4) | 0.67 |
| **LV Diastolic Function** | 78 | 66 (84.6) | 1 (1.3) | 11 (14.1) | 0.72 |
| **LV Wall Thickness** | 58 | 33 (56.9) | 0 (0) | 25 (43.1) | 0.60 |

**Abbreviations:** AVA Cont VTI, Aortic Valve Area Calculated by Velocity Time Integral ; AVA Index, Aortic Valve Area Index; AV Mn Grad, Aortic Valve Mean Gradient; AIPHT, Aortic Insufficiency Pressure Half-Time; AV Pk Vel, Aortic Valve Peak Velocity; AV Regurgitation, Aortic Valve Regurgitation; AV Stenosis, Aortic Valve Stenosis; AV Structure, Aortic Valve Structure; GLS, Global Longitudinal Strain; IVSd, Interventricular Septum Thickness; ; LV Diastolic Function, Left Ventricular Diastolic Function; LVOT Pk Grad, Left Ventricular Outflow Tract Peak Gradient; LVOT Pk Vel, Left Ventricular Outflow Tract Peak Velocity; LV Wall Thickness, Left Ventricular Wall Thickness; MV Regurgitation, Mitral Valve Regurgitation; MV Stenosis, Mitral Valve Stenosis; MV Structure, Mitral Valve Structure.

### External Validation: Model performance in MIMIC-III and MIMIC-IV TTE Reports

In 100 TTE reports from the MIMIC-III dataset, HeartDx-LM also demonstrated high accuracy in extracting structured clinical data. The model successfully extracted 615 out of 707 available values correctly, achieving an overall accuracy rate of 86.9% and Cohen’s Kappa of 0.90. This included an accuracy of 72.4% for continuous variables (113/156) and 91.1% for categorical variables (502/551).

There were 12 (1.7%) values inaccurately extracted and 80 (11.3%) failed extractions, mainly in the qualitative labels. The inaccurate values were most frequent for AV structure (4/95), followed by AV regurgitation (2/91), AV stenosis (2/61), and MV regurgitation (1/93). The model failed to extract 80 (11.3%) values in the reports across all 18 variables. External validation in the MIMIC III dataset also demonstrated high accuracy, with the model achieving over 90% accuracy for key continuous and categorical clinical variables (e.g. EF: 97.8% [90/92], AV structure: 94.7% [90/95], MV regurgitation: 97.8% [88/90], and LV wall thickness: 92.7% [76/82]; **Table 4**).

**Table 4:** Model Performance Evaluation of HeartDX-LM on MIMIC-III TTE Reports.

| Metrics | Available Values | Correct: N (%)<br>(Extracted value matches original value) | Incorrect: N (%)<br>(Extracted value differs from original value) | Failed: N (%)<br>(Value not extracted) | Cohen's Kappa |
| --- | --- | --- | --- | --- | --- |
| Column |  |  |  |  |  |
| All variables | 707 | 615 (86.9) | 12 (1.7) | 80 (11.3) | 0.90 |
| AIPHT | 0 | - | - | - | - |
| AVA Cont VTI | 7 | 0 (0) | 0 (0) | 7 (100) | 0.48 |
| AVA Index | 0 | - | - | - | - |
| AV Mn Grad | 3 | 3 (100) | 0 (0) | 0 (0) | 1.0 |
| AV Pk Vel | 0 | - | - | - | - |
| Ejection Fraction | 92 | 90 (97.8) | 0 (0) | 2 (2.2) | 0.88 |
| GLS% | 0 | - | - | - | - |
| IVSd | 1 | 1 (100) | 0 (0) | 0 (0) | 1.0 |
| LVOT Pk Grad | 53 | 19 (35.8) | 0 (0) | 34 (64.2) | 0.50 |
| LVOT Pk Vel | 0 | - | - | - | - |
| AV Structure | 95 | 90 (94.7) | 4 (4.2) | 1 (1.1) | 0.65 |
| AV Stenosis | 61 | 51 (83.6) | 2 (3.3) | 8 (13.1) | 0.81 |
| AV Regurgitation | 91 | 81 (89) | 2 (2.2) | 8 (8.8) | 0.61 |
| MV Structure | 87 | 85 (97.7) | 2 (2.3) | 0 (0) | 0.92 |
| MV Stenosis | 21 | 13 (61.9) | 0 (0) | 8 (38.1) | 0.77 |
| MV Regurgitation | 90 | 88 (97.8) | 0 (0) | 2 (2.2) | 0.90 |
| LV Diastolic Function | 24 | 18 (75) | 2 (8.3) | 4 (16.7) | 0.84 |
| LV Wall Thickness | 82 | 76 (92.7) | 0 (0) | 6 (7.3) | 0.83 |

**Abbreviations:** AVA Cont VTI, Aortic Valve Area Calculated by Velocity Time Integral ; AVA Index, Aortic Valve Area Index; AV Mn Grad, Aortic Valve Mean Gradient; AIPHT, Aortic Insufficiency Pressure Half-Time; AV Pk Vel, Aortic Valve Peak Velocity; AV Regurgitation, Aortic Valve Regurgitation; AV Stenosis, Aortic Valve Stenosis; AV Structure, Aortic Valve Structure; GLS, Global Longitudinal Strain; IVSd, Interventricular Septum Thickness; ; LV Diastolic Function, Left Ventricular Diastolic Function; LVOT Pk Grad, Left Ventricular Outflow Tract Peak Gradient; LVOT Pk Vel, Left Ventricular Outflow Tract Peak Velocity; LV Wall Thickness, Left Ventricular Wall Thickness; MV Regurgitation, Mitral Valve Regurgitation; MV Stenosis, Mitral Valve Stenosis; MV Structure, Mitral Valve Structure.

In the MIMIC-IV dataset, the model successfully extracted 201 out of 220 available values, achieving an overall accuracy rate of 91.3% and Cohen’s Kappa value of 0.95. This included an accuracy of 97.8% (44/45) for continuous variables and 89.7% (157/175) for categorical variables. The model extracted 2 (0.9%) incorrect values, 1 out of 45 values of EF and 1 out of 42 values MV regurgitation. Additionally, the model failed to extract 17 (7.7%) values present in the reports across the 18 variables. Values of accuracy for specific labels can be found in **Table 5**. The performance of HeartDX-LM across all 4 datasets is summarized in **Figure 3**.

**Table 5:** Model Performance Evaluation of HeartDX-LM on MIMIC-IV Reports.

| Metrics | Available Values | Correct: N (%)<br>(Extracted value matches original value) | Incorrect: N (%)<br>(Extracted value differs from original value) | Failed: N (%)<br>(Value not extracted) | Cohen's Kappa |
| --- | --- | --- | --- | --- | --- |
| Column |  |  |  |  |  |
| **All variables** | 220 | 201 (91.3) | 2 (0.9) | 17 (7.7) | 0.95 |
| **AIPHT** | 0 | - | - | - | - |
| **AVA Cont VTI** | 0 | - | - | - | - |
| **AVA Index** | 0 | - | - | - | - |
| **AV Mn Grad** | 0 | - | - | - | - |
| **AV Pk Vel** | 0 | - | - | - | - |
| **Ejection Fraction** | 45 | 44 (97.8) | 1 (2.2) | 0 (0) | 0.98 |
| **GLS%** | 0 | - | - | - | - |
| **IVSd** | 0 | - | - | - | - |
| **LVOT Pk Grad** | 0 | - | - | - | - |
| **LVOT Pk Vel** | 0 | - | - | - | - |
| **AV Structure** | 25 | 22 (88) | 0 (0) | 3 (12) | 0.92 |
| **AV Stenosis** | 24 | 23 (95.8) | 0 (0) | 1 (4.2) | 0.97 |
| **AV Regurgitation** | 34 | 33 (97) | 0 (0) | 1 (3) | 0.98 |
| **MV Structure** | 32 | 27 (84.4) | 0 (0) | 5 (15.6) | 0.89 |
| **MV Stenosis** | 3 | 2 (66.7) | 0 (0) | 1 (33.3) | 0.83 |
| **MV Regurgitation** | 43 | 38 (88.4) | 1 (1.1) | 4 (9.3) | 0.90 |
| **LV Diastolic Function** | 4 | 4 (100) | 0 (0) | 0 (0) | 1.0 |
| **LV Wall Thickness** | 10 | 8 (80) | 0 (0) | 2 (20) | 0.89 |

**Abbreviations:** AVA Cont VTI, Aortic Valve Area Calculated by Velocity Time Integral ; AVA Index, Aortic Valve Area Index; AV Mn Grad, Aortic Valve Mean Gradient; AIPHT, Aortic Insufficiency Pressure Half-Time; AV Pk Vel, Aortic Valve Peak Velocity; AV Regurgitation, Aortic Valve Regurgitation; AV Stenosis, Aortic Valve Stenosis; AV Structure, Aortic Valve Structure; GLS, Global Longitudinal Strain; IVSd, Interventricular Septum Thickness; ; LV Diastolic Function, Left Ventricular Diastolic Function; LVOT Pk Grad, Left Ventricular Outflow Tract Peak Gradient; LVOT Pk Vel, Left Ventricular Outflow Tract Peak Velocity; LV Wall Thickness, Left Ventricular Wall Thickness; MV Regurgitation, Mitral Valve Regurgitation; MV Stenosis, Mitral Valve Stenosis; MV Structure, Mitral Valve Structure.

### Data Volume for Model Fine-tuning

In our evaluation of the data threshold necessary for model development, we analyzed the accuracy of our models as a function of progressively larger number of reports used for fine-tuning. The Llama2-13b models finetuned using 100 and 200 reports had accuracies of 13.5% and 85.7%, respectively. The accuracies increased to 97.8%, 98.2%, and 98.8 with the use of 500, 1000, and 2000 TTE reports, respectively (**Figure 2**). A minimum of 500 reports were necessary to achieve our pre-specified accuracy benchmark of 95%, with accuracy plateauing beyond this point.

**Figure 2.**
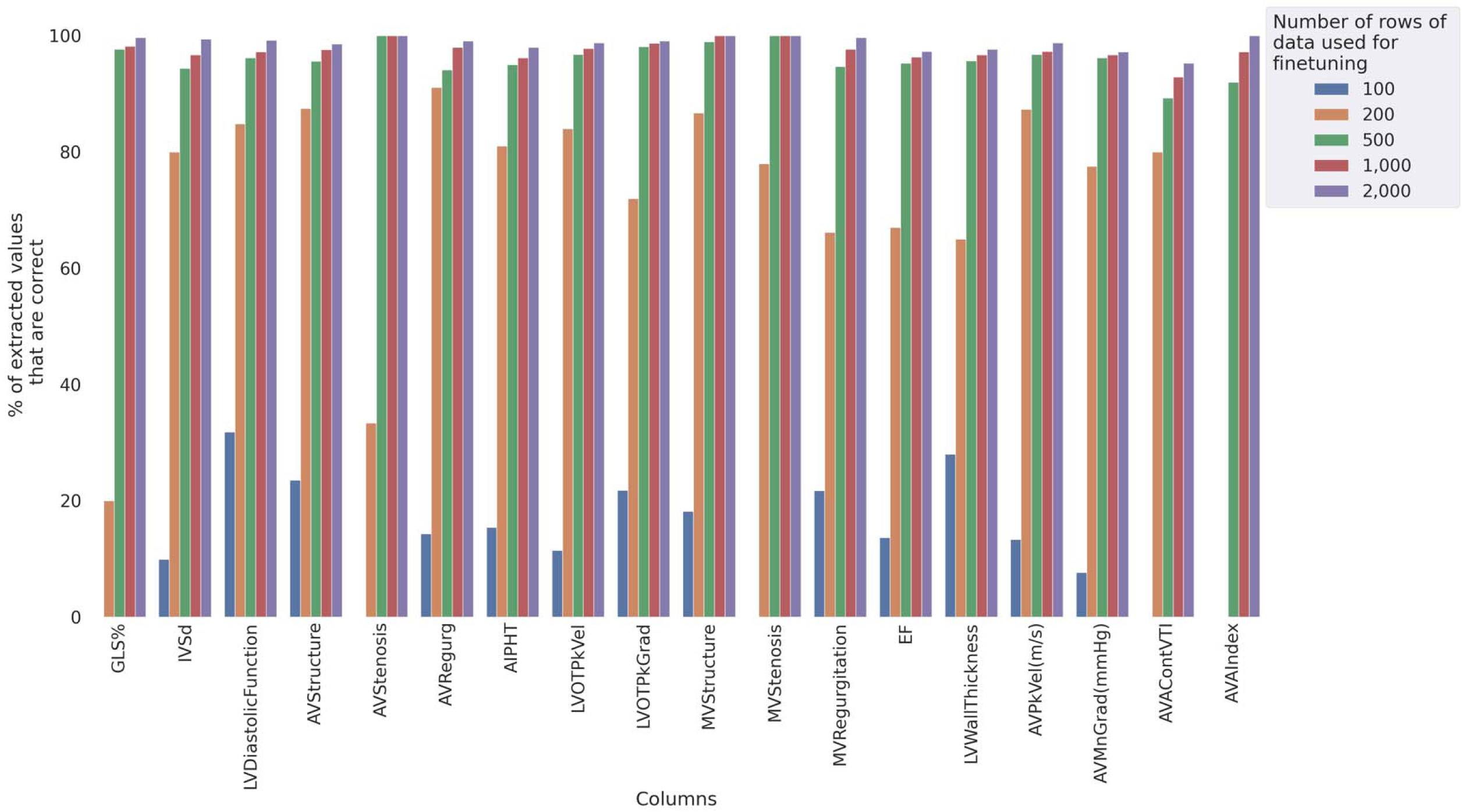
Performance of models fine-tuned with varying number of paired unstructured reports and structured tables for tabulation of clinical variables from unstructured reports. **Abbreviations**: AVA Cont VTI, Aortic Valve Area Calculated by Velocity Time Integral; AVA Index, Aortic Valve Area Index; AV Mn Grad, Aortic Valve Mean Gradient; AIPHT, Aortic Insufficiency Pressure Half-Time; AV Pk Vel, Aortic Valve Peak Velocity; AV Regurgitation, Aortic Valve Regurgitation; AV Stenosis, Aortic Valve Stenosis; AV Structure, Aortic Valve Structure; GLS, Global Longitudinal Strain; IVSd, Interventricular Septum Thickness; LV Diastolic Function, Left Ventricular Diastolic Function; LVOT Pk Grad, Left Ventricular Outflow Tract Peak Gradient; LVOT Pk Vel, Left Ventricular Outflow Tract Peak Velocity; LV Wall Thickness, Left Ventricular Wall Thickness; MV Regurgitation, Mitral Valve Regurgitation; MV Stenosis, Mitral Valve Stenosis; MV Structure, Mitral Valve Structure.

**Figure 3.**
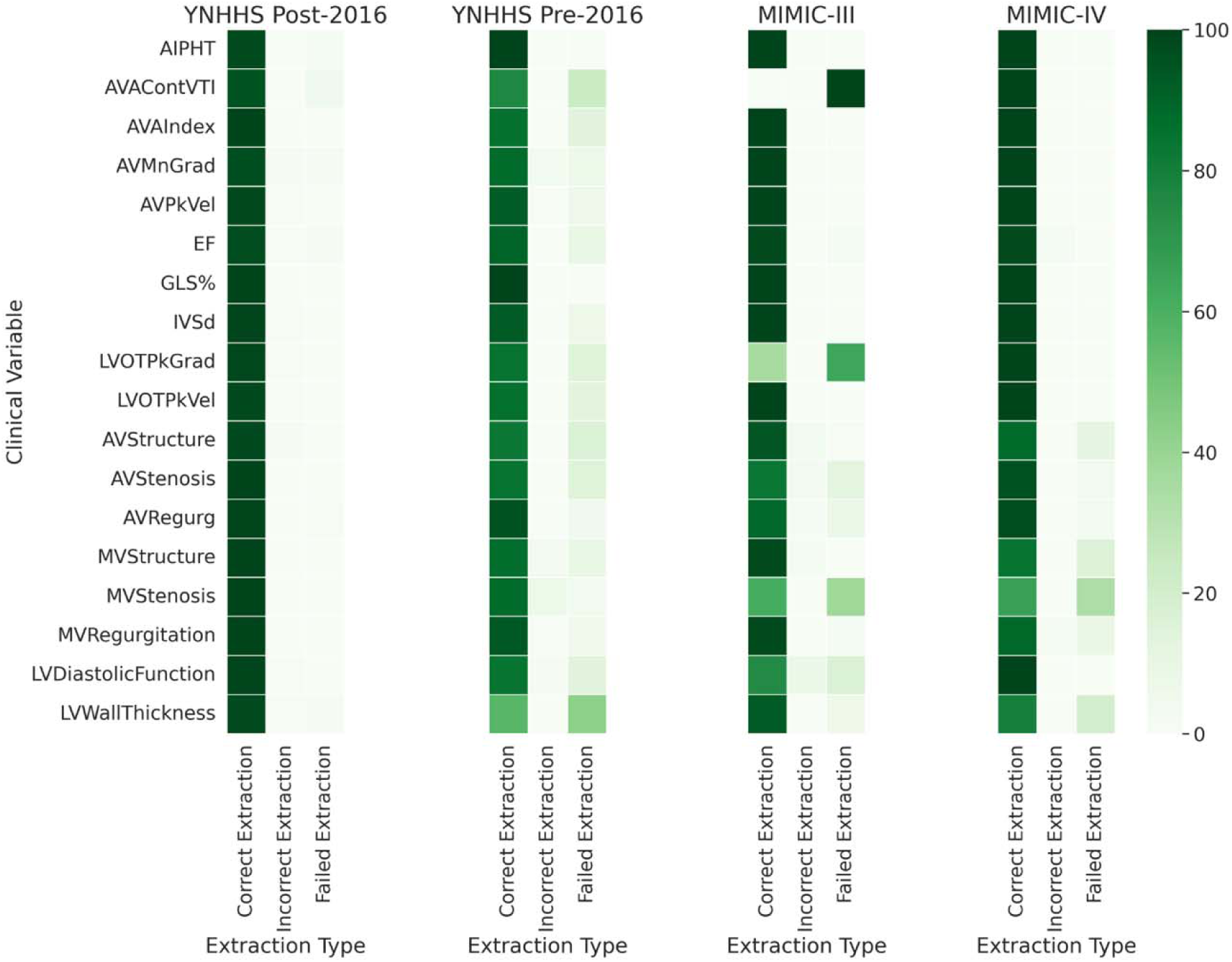
Accuracy of HeartDX-LM for Label Extraction Across the Four Datasets. Abbreviations: AVA Cont VTI, Aortic Valve Area Calculated by Velocity Time Integral; AVA Index, Aortic Valve Area Index; AV Mn Grad, Aortic Valve Mean Gradient; AIPHT, Aortic Insufficiency Pressure Half-Time; AV Pk Vel, Aortic Valve Peak Velocity; AV Regurgitation, Aortic Valve Regurgitation; AV Stenosis, Aortic Valve Stenosis; AV Structure, Aortic Valve Structure; GLS, Global Longitudinal Strain; IVSd, Interventricular Septum Thickness; LV Diastolic Function, Left Ventricular Diastolic Function; LVOT Pk Grad, Left Ventricular Outflow Tract Peak Gradient; LVOT Pk Vel, Left Ventricular Outflow Tract Peak Velocity; LV Wall Thickness, Left Ventricular Wall Thickness; MV Regurgitation, Mitral Valve Regurgitation; MV Stenosis, Mitral Valve Stenosis; MV Structure, Mitral Valve Structure.

## DISCUSSION

We developed and validated HeartDx-LM, an innovative strategy to extract structured clinical data from unstructured clinical reports. This novel strategy leverages the output of an LLM to train a smaller, lightweight model, eliminating the need for high computational capacity in the final deployment. HeartDx-LM demonstrated robust performance in digitizing TTE reports across varying reporting formats from geographically and temporally distinct data sources and was able to successfully extract qualitative and quantitative clinical labels with high accuracy. The model’s adaptability and extensibility enable its potential deployment in diverse and low-resource clinical settings and applicability to other diagnostic reports. Furthermore, our research determined the minimum threshold for the number of TTE reports required for fine-tuning models for optimally balanced accuracy and computational resources, providing valuable guidance for future model development.

Prior models to digitize TTE reports predominantly relied on rule-based or keyword-based NLP models.^21–23^ For example, early studies have used specific keywords and predefined rules to analyze echocardiography and radiology reports without considering variations in reporting formats and dynamic changes in clinical parameters.^24–26^ Moreover, these methods predominantly focus on extracting a few specific clinical labels, such as low EF, and often fail to capture the full spectrum of clinically relevant labels needed for broader applications, healthcare decision-making, and planning.^27^

In contrast, HeartDx-LM, was engineered to extract multiple qualitative and quantitative clinical labels. This comprehensive extraction capability enhances the model’s utility in clinical practice as it can be scaled to similar domains, where diagnostic information is captured in unstructured reports. This innovative two-step approach to digitizing entire reports is an alternative for generating training sets for smaller LLMs, reducing the need for extensive manual annotation and the reliance on high computational power. Since most TTE reports are stored as unstructured text, this approach can significantly expand our dataset for new model training, and enable access to diverse settings, including those with limited technological infrastructure, with potential use for cross-setting electronic clinical quality measures.^2,28–31^

Our study has limitations that deserve consideration. Notably, the performance of our models showed variability across different clinical fields, especially when certain domain-specific terms were reported differently across different datasets. Nonetheless, the overall and field-wise performance was acceptable across all external sites. Additionally, the computational resources required for finetuning LLMs may pose practical constraints in real-world healthcare settings. However, the deployment of the lightweight finetuned model, that we have also publicly released on HuggingFace, does not require intensive computational resources and can be used for transformation of unstructured reports into tabular datasets. Finally, while our study underscores the potential in using LLMs for the automated extraction of structured clinical information from unstructured narratives in EHR, future research should prioritize enhancing the interpretability of LLM-based models. This can be achieved by delving into the contextual analysis of clinical notes and refining the model’s ability to discern subtle nuances in medical language to further optimize the performance and generalizability of LLM-based approaches.

## CONCLUSION

We developed a novel method using paired large and moderate-sized LLMs to automate the extraction of unstructured echocardiographic reports into tabular datasets. Our approach represents a scalable strategy that transforms unstructured reports into computable elements that can be leveraged to improve cardiovascular care quality and enable research.

## Funding

Dr. Khera was supported by the National Heart, Lung, and Blood Institute of the National Institutes of Health (under awards R01AG089981, R01HL167858, and K23HL153775) and the Doris Duke Charitable Foundation (under award 2022060). Dr. Oikonomou was supported by the National Heart, Lung, and Blood Institute of the National Institutes of Health (under award F32HL170592). The funders had no role in the design and conduct of the study; collection, management, analysis, and interpretation of the data; preparation, review, or approval of the manuscript; and decision to submit the manuscript for publication.

## Conflict of Interest

Dr. Khera is an Associate Editor of JAMA and is a co-founder of Ensight-AI. Dr. Khera receives support from the National Heart, Lung, and Blood Institute of the National Institutes of Health (under awards R01HL167858 and K23HL153775) and the Doris Duke Charitable Foundation (under award 2022060). He receives support from the Blavatnik Foundation through the Blavatnik Fund for Innovation at Yale. He also receives research support, through Yale, from Bristol-Myers Squibb, BridgeBio, and Novo Nordisk. In addition to 63/346,610, Dr. Khera is a coinventor of U.S. Pending Patent Applications WO2023230345A1, US20220336048A1, 63/484,426, 63/508,315, 63/580,137, 63/619,241, 63/346,610, 63/562,335 and 18/813,882. Dr. Khera, Dr. Oikonomou and Mr. Vasisht Shankar are co-inventors of the US patent application 63/606,203. Dr. Khera and Dr. Oikonomou are co-founders of Evidence2Health, a precision health platform to improve evidence-based cardiovascular care. Dr. Oikonomou receives support from the National Heart, Lung, and Blood Institute of the National Institutes of Health (under award F32HL170592). He is a co-inventor of the U.S. Patent Applications 18/813,882, 17/720,068, 63/619,241, 63/177,117, 63/580,137, 63/606,203, 63/562,335, US11948230B2, US20210374951A1. He has been a consultant for Caristo Diagnostics Ltd and Ensight-AI Inc, and has received royalty fees from technology licensed through the University of Oxford.. Mr. Vasisht Shankar works as a data scientist at Evidence2Health (outside the current work). Dr. Nadkarni is a founder of Renalytix, Pensieve, and Verici and provides consultancy services to AstraZeneca, Reata, Renalytix, and Pensieve. He also has equity in Renalytix, Pensieve, and Verici.

## Data Availability

The dataset cannot be made publicly available because they are electronic health records. Sharing this data externally without proper consent could compromise patient privacy and would violate the Institutional Review Board’s approval for the study.

## Code Availability

The code for the study is available from the authors upon request.

