## Supplementary material for "Automated Transformation of Unstructured Cardiovascular Diagnostic Reports into Structured Datasets Using Sequentially Deployed Large Language Models": Online Supplement

**Table S1. Examples of de-identified TTE reports from the 4 datasets.**

| **Dataset** | **De-identified TTE Report** |
| --- | --- |
| YNHHS post-2016 | Findings: * Moderately decreased left ventricular systolic function. Mild concentric left ventricular hypertrophy. LVEF estimated by visual assessment is between 30-35%. * The entire apex, the mid anterolateral, and mid inferoseptal segments are akinetic. * Normal right ventricular cavity size, wall thickness, and systolic function. * The mid inferior, mid anteroseptal, and mid inferolateral segments are hypokinetic. * The mitral valve appears thickened. There is moderate mitral valve regurgitation. * There is mild tricuspid valve regurgitation. The pulmonary artery systolic pressure is 40.00 mmHg. * There is mild aortic valve regurgitation. * There is no evidence of pericardial effusion. Conclusion: Left Ventricle Normal left ventricular cavity size. Moderately decreased left ventricular systolic function. Mild concentric left ventricular hypertrophy. LVEF estimated by visual assessment is between 30-35%. Wall Motion Rest Echo Findings The mid inferior, mid anteroseptal, and mid inferolateral segments are hypokinetic. The entire apex, the mid anterolateral, and mid inferoseptal segments are akinetic. All other scored wall segments showed normal motion. Right Ventricle Normal right ventricular cavity size, wall thickness, and systolic function. There is an ICD/Pacer wire seen in the right ventricle. Atria Mild biatrial enlargement. Aortic Valve Normal aortic valve structure and function. There is mild aortic valve regurgitation. Mitral Valve The mitral valve appears thickened. There is moderate mitral valve regurgitation. There is no mitral valve stenosis. Tricuspid Valve Normal tricuspid valve structure. There is mild tricuspid valve regurgitation. The pulmonary artery systolic pressure is 40.00 mmHg. Pulmonic Valve The pulmonic valve is normal. There is trace to mild pulmonic valve regurgitation. Great Vessels Dilatation of the sinuses of Valsalva measuring 3.80 cm and dilatation of the ascending aorta measuring 4.20 cm. Venous The inferior vena cava was not well visualized. Pericardium/Pleural There is no evidence of pericardial effusion. Prior Study Comparison No significant image changes compared with prior study. |
| YNHHS pre-2016 | Transthoracic Echo Report Ht: 175 cm Wt: 82 kg BSA: 2.01 m^2 BP: 136 / 77 HR: 72 Indications: Abnormal EKG Procedure: COMPLETE 2D, DOPPLER AND COLOR Agitated Saline: Yes Contrast: None 3D Imaging of: Left Ventricle Technical Quality: Good IMPRESSION Normal left and right ventricular size and systolic function. Estimated ejection fraction estimated by 3D volumetric analysis is 66 Normal diastolic function. Mild left atrial dilatation. Early right-to-left shunt by agitated saline (bubble) study, suggesting intracardiac shunt. Aneurysmal interatrial septum. No signficant valvular abnormalities. No prior studies for comparison. FINDINGS LEFT VENTRICLE: Normal left ventricular size. Normal wall thickness. Normal systolic function. No regional wall motion abnormalities. Estimated ejection fraction estimated by 3D volumetric analysis is 66 %. Normal diastolic function. RIGHT VENTRICLE: Normal right ventricular size. Normal systolic function. Estimated right ventricular systolic pressure is 30 mmHg. ATRIA: Mild left atrial dilatation. Normal right atrial size. Early right- to-left shunt by agitated saline (bubble) study, suggesting intracardiac shunt. Aneurysmal interatrial septum. MITRAL VALVE: Normal appearing mitral valve leaflets. No significant mitral regurgitation. AORTIC VALVE: Trileaflet aortic valve. No aortic stenosis. No significant aortic regurgitation. TRICUSPID VALVE: Structurally normal tricuspid valve leaflets. Mild tricuspid regurgitation. PULMONARY VALVE: Structurally normal pulmonic valve leaflets. No pulmonary regurgitation. AORTA: Normal visualized portions of the aorta. INFERIOR VENA CAVA: Normal inferior vena cava. PERICARDIUM: No significant pericardial effusion. MEASUREMENTS 2D ECHO LV Diastolic Diameter PLAX 5.3 cm (4.1 - 5.7) LV Systolic Diameter PLAX 3.4 cm (2.0 - 4.0 IVS Diastolic Thickness 0.7 cm (0.6 - 1.0 ) LVPW Diastolic Thickness 0.9 cm (0.6 - 1.0) LA Systolic Diameter LX 3.8 cm (3.0 - 4.0) LA Volume MOD BP Index 38.4 ml/m? (16 - 28 cm?/m?) Aortic Root Diameter 2.9 cm (2.7-4.1) DOPPLER LVOT Peak Velocity 1.3 m/s AV Peak Velocity 1.6 m/s (1.0-1.7 m/s) TR Peak Velocity 2.3 m/s TR Peak Gradient 21.4 mmHg (< 25 mmHg) Mitral E Point Velocity 1.1 m/s (0.6-1.3) Mitral A Point Velocity 0.7 m/s (0.3 -0.6) Mitral E to A Ratio 1.6 MV Deceleration Time 217.0 ms LV E' Lateral Velocity 12.2 cm/s (>=10cm/s) LV E' Septal Velocity 12.4 cm/s (>=8cm/s) Mitral E to LV E' Lateral Ratio 8.8 (=<8) Mitral E to LV E' Septal Ratio 8.6 (=<8) |
| MIMIC-III | PATIENT/TEST INFORMATION: Indication: Endocarditis. Height: (in) 74 Weight (lb): 165 BSA (m2): 2.00 m2 BP (mm Hg): 102/72 HR (bpm): 86 Status: Inpatient Date/Time: [**2102-6-14**] at 13:30 Test: Portable TTE (Complete) Doppler: Full Doppler and color Doppler Contrast: None Technical Quality: Adequate INTERPRETATION: Findings: LEFT ATRIUM: Normal LA and RA cavity sizes. LEFT VENTRICLE: Normal LV wall thickness, cavity size and regional/global systolic function (LVEF >55%). No resting LVOT gradient. RIGHT VENTRICLE: Normal RV chamber size and free wall motion. AORTA: Moderately dilated aortic sinus. Normal ascending aorta diameter. Normal aortic arch diameter. No 2D or Doppler evidence of distal arch coarctation. AORTIC VALVE: Normal aortic valve leaflets (3). No AS. No AR. No masses or vegetations on aortic valve. MITRAL VALVE: Normal mitral valve leaflets with trivial MR. No MVP. No mass or vegetation on mitral valve. Normal mitral valve supporting structures. TRICUSPID VALVE: Normal tricuspid valve leaflets with trivial TR. Normal PA systolic pressure. PULMONIC VALVE/PULMONARY ARTERY: Normal pulmonic valve leaflet. No PS. Physiologic PR. PERICARDIUM: No pericardial effusion. Conclusions: The left atrium and right atrium are normal in cavity size. Left ventricular wall thickness, cavity size and regional/global systolic function are normal (LVEF >55%) Right ventricular chamber size and free wall motion are normal. The aortic root is moderately dilated at the sinus level. The aortic valve leaflets (3) appear structurally normal with good leaflet excursion and no aortic regurgitation. No masses or vegetations are seen on the aortic valve. The mitral valve appears structurally normal with trivial mitral regurgitation. There is no mitral valve prolapse. No mass or vegetation is seen on the mitral valve. The estimated pulmonary artery systolic pressure is normal. There is no pericardial effusion. IMPRESSION: No vegetation or abscess seen. Normal global and regional biventricular systolic function. No pulmonary hypertension or significant valvular disease seen. If clinically suggested, the absence of a vegetation by 2D echocardiography does not exclude endocarditis. |
| MIMIC-IV | ECHO The left atrium is mildly dilated. Left ventricular wall thicknesses and cavity size are normal. There is mild regional left ventricular systolic dysfunction with focal apical hypokinesis. The remaining segments contract normally (LVEF = 55%). No masses or thrombi are seen in the left ventricle. Right ventricular chamber size and free wall motion are normal. There are three aortic valve leaflets. There is mild aortic valve stenosis (valve area 1.7cm2). Mild (1+) aortic regurgitation is seen. The mitral valve leaflets are mildly thickened. Trivial mitral regurgitation is seen. The pulmonary artery systolic pressure could not be determined. There is a trivial/physiologic pericardial effusion. Normal left ventricular cavity size with mild regional systolic dysfunction most c/w CAD (distal LAD distribution). Mild aortic valve stenosis. Mild aortic regurgitation. |

**Table S2. Variations in free text reporting across echocardiographers in the pre-2016 YNHHS reports**

| **Variation** | **De-identified TTE Report** |
| --- | --- |
| **1** | Transthoracic Echo Report: Ht: 175 cm, Wt: 82 kg, BSA: 2.01 m^2, BP: 136/77, HR: 72, Indications: Abnormal EKG, Procedure: COMPLETE 2D, DOPPLER AND COLOR, Agitated Saline: Yes, Contrast: None, 3D Imaging of: Left Ventricle, Technical Quality: Good. IMPRESSION: Normal left and right ventricular size and systolic function. Estimated ejection fraction by 3D volumetric analysis is 66%. Normal diastolic function. Mild left atrial dilatation. Early right-to-left shunt by agitated saline (bubble) study, suggesting intracardiac shunt. Aneurysmal interatrial septum. No significant valvular abnormalities. No prior studies for comparison. FINDINGS: LEFT VENTRICLE: Normal left ventricular size. Normal wall thickness. Normal systolic function. No regional wall motion abnormalities. Estimated ejection fraction by 3D volumetric analysis is 66%. Normal diastolic function. RIGHT VENTRICLE: Normal right ventricular size. Normal systolic function. Estimated right ventricular systolic pressure is 30 mmHg. ATRIA: Mild left atrial dilatation. Normal right atrial size. Early right-to-left shunt by agitated saline (bubble) study, suggesting intracardiac shunt. Aneurysmal interatrial septum. MITRAL VALVE: Normal appearing mitral valve leaflets. No significant mitral regurgitation. AORTIC VALVE: Trileaflet aortic valve. No aortic stenosis. No significant aortic regurgitation. TRICUSPID VALVE: Structurally normal tricuspid valve leaflets. Mild tricuspid regurgitation. PULMONARY VALVE: Structurally normal pulmonic valve leaflets. No pulmonary regurgitation. AORTA: Normal visualized portions of the aorta. INFERIOR VENA CAVA: Normal inferior vena cava. PERICARDIUM: No significant pericardial effusion. MEASUREMENTS: 2D ECHO: LV Diastolic Diameter PLAX 5.3 cm (4.1 - 5.7), LV Systolic Diameter PLAX 3.4 cm (2.0 - 4.0), IVS Diastolic Thickness 0.7 cm (0.6 - 1.0), LVPW Diastolic Thickness 0.9 cm (0.6 - 1.0), LA Systolic Diameter LX 3.8 cm (3.0 - 4.0), LA Volume MOD BP Index 38.4 ml/m² (16 - 28 cm²/m²), Aortic Root Diameter 2.9 cm (2.7 - 4.1). DOPPLER: LVOT Peak Velocity 1.3 m/s, AV Peak Velocity 1.6 m/s (1.0 - 1.7 m/s), TR Peak Velocity 2.3 m/s, TR Peak Gradient 21.4 mmHg (< 25 mmHg), Mitral E Point Velocity 1.1 m/s (0.6 - 1.3), Mitral A Point Velocity 0.7 m/s (0.3 - 0.6), Mitral E to A Ratio 1.6, MV Deceleration Time 217.0 ms, LV E' Lateral Velocity 12.2 cm/s (>= 10 cm/s), LV E' Septal Velocity 12.4 cm/s (>= 8 cm/s), Mitral E to LV E' Lateral Ratio 8.8 (=< 8), Mitral E to LV E' Septal Ratio 8.6 (=< 8). |
| **2** | **Diagnosis:** Aortic stenosis **Reason for Test:** Evaluate LV function and valves **Study:** 2D & M-mode: X Doppler X **Ht:** 5'1" **Wt:** 118 lbs. **BSA:** 1.5 m² **BP:** 130/70 **CONCLUSIONS:** Severe aortic stenosis. Peak gradient 80 mmHg/mean gradient 16 mmHg. Estimated valve area 0.71 sq cm. Mild concentric left ventricular hypertrophy. Ejection fraction 60% with diastolic dysfunction and suggestion of elevated filling pressures. Mitral annular calcification interferes with this measurement. Moderate mitral annular calcification with moderate MR. Moderate left atrial enlargement. Normally contractile right ventricle. No estimate of RVSP possible. Comparing this to the prior study there is no meaningful change. **ECHOCARDIOGRAPHIC FINDINGS** Aortic root, ascending aorta and arch: Normal caliber. Aortic valve: Calcified, trileaflet, severe stenosis. Left atrium: Moderately enlarged. Mitral valve: Fibrocalcific change. No prolapse. Mitral valve annulus: Calcified. Tricuspid valve: Opens normally. No TR. Pulmonic valve: Not well seen. Left ventricle: Mild concentric left ventricular hypertrophy. Normal ejection fraction 60%. Diastolic dysfunction with estimated elevated filling pressures. Normal regional wall motion. Right ventricle: Normal right ventricular chamber size and systolic function. Right atrium: Normal size. Inferior vena cava: Normal caliber; normal phasic variation. Pericardium: No pericardial effusion. **DOPPLER FINDINGS** CW, PW & CF: Aortic valve: Peak velocity: 4.5 m/sec. Peak instantaneous gradient: 80 mm Hg. Mean gradient: 60 mm Hg. Estimated valve area (continuity equation): 0.71 cm². Mitral valve: Valve area (Doppler): cm². Valve area (planimetry): cm². Mean transvalvular gradient: 2 mm Hg. Est. PA systolic/diastolic pressure: mmHg. Regurgitation Detected: Mitral: Moderate. Aortic: Mild. Tricuspid: None. Pulmonic: None. Diastolic function DTI: Septal E: 0.96. E1: 0.03. E/E1: 32. Lateral E: 0.96. E1: 0.05. E/E1: 21. A Dur: 180 msec. Deceleration Time: 295 msec. IVRT: 1:30 msec. **ECHOCARDIOGRAPHIC MEASUREMENTS** Aortic root: Measured 2.8 cm. Index: 1.9 cm/m². Normal <2.2 cm/m². Ascending Aorta: Measured 2.7 cm. Index: 1.8 cm/m². Normal <2.2 cm/m². Aortic arch: Measured 2.5 cm. Index: 1.7 cm/m². Normal <2.2 cm/m². Aortic valve excursion: Measured cm. Normal 1.6 - 2.6. Left atrium: Measured 3.7 cm. Index: 2.5 cm/m². Normal <2.1 cm/m². Left atrial volume: 49 ml/m². Normal < 29 ml/m². LV septum diastolic thickness: 12 mm. 6-10 mm. LV posterior wall diastolic thickness: 11 mm. 6-10 mm. Left ventricular dimension: End diastolic size: 43 mm (42-59 mm). End systolic size: 32 mm. EDV: 82 ml (67-155 ml). EDVi: 55 ml/m² (35-75 ml/m²). ESV: 42 ml (22-58 ml). ESVi: 28 ml/m² (12- ml/m²). SV: ml. SVi: 27 ml/m². Visual LVEF: 60%. Modified Simpson's LVEF: 61%. **CC:** |
| **3** | The findings indicate hyperdynamic left ventricular systolic function with mild LVOT obstruction, reaching a peak gradient of 15 mmHg with Valsalva. Aortic sclerosis is present without stenosis, and there is mild aortic valve regurgitation. The left ventricle shows normal cavity size and severe concentric hypertrophy, with moderate diastolic dysfunction consistent with increased left atrial pressure. The right ventricle has normal cavity size, wall thickness, systolic function, and wall motion. The left atrium is mildly dilated, while the right atrium is normal in size. The aortic valve is trileaflet with mild calcification and sclerosis, but without stenosis, and there is mild aortic valve regurgitation with a pressure half-time of 750.0773569 msec. The mitral valve shows normal leaflets with trace regurgitation, and the tricuspid valve is normal with trace regurgitation. The measurements include a normal aortic valve cusp size of 2.07 cm/m², left ventricular internal diameter in diastole (LVIDd) of 4.5 cm, and end-diastolic volume (EDV) of 92.00 ml. The ejection fraction (EF) is 69% (4-chamber view), 78% (2-chamber view), and 74% (biplane). The mitral valve peak E wave velocity is 0.7 m/sec with a deceleration time of 260 ms. |
| **4** | **Impression:** Moderate to severe mitral regurgitation with severe left atrial enlargement and mild left ventricular enlargement - this may be somewhat improved compared to prior studies. Borderline left ventricular ejection fraction at 50% with paced asyneresis, apical hypokinesis, and possible inferolateral hypokinesis. Biologic aortic valve replacement with normal function. Much improved mild tricuspid regurgitation with significant improvement in pulmonary artery pressure from mid-70s to about 40. Mild right-sided enlargement with borderline systolic function. Pacing wires noted in the right heart.  **Echocardiographic findings:** The acoustic windows were average. The left atrium was markedly enlarged. The left ventricle was mildly enlarged. There was no hypertrophy. Regional wall motion was abnormal with paced asyneresis and apical/inferolateral apical hypokinesis. The ejection fraction was borderline preserved and visually estimated at 50%. Diastolic parameters suggested elevated filling pressures. The visualized thoracic aorta was not dilated. The right atrium was enlarged. The right ventricle was mildly enlarged. Right ventricular systolic function was borderline normal. The pulmonary artery was not dilated. There was no significant pericardial effusion. The inferior vena cava was prominent. The aortic valve was replaced with a bioprosthetic valve. The valve was well seated and normally functioning. There was no significant stenosis and no insufficiency. The mitral valve was sclerotic without myxomatous or rheumatic changes. There was moderate annular sclerosis. There was moderate in some views and severe in other views regurgitation. There was mild functional stenosis from the degree of regurgitation. The pulmonic valve appeared normal on limited views with trace regurgitation. The tricuspid valve was anatomically normal with mild regurgitation. There was no significant Doppler evidence for intracardiac shunting on the present study.  **Doppler Findings CW, PW, and CF:** Regurgitation Detected: Mitral: Moderate to severe, Aortic: None significant, Tricuspid: Mild, Pulmonic: Trace. Aortic valve: Peak velocity: 1.9 m/sec, Peak instantaneous gradient: 15 mm Hg, Mean gradient: 8 mm Hg, Estimated valve area (continuity equation): 1.2 cm², Dimensionless orifice index: 0.259/0.451 = 0.57. Mitral valve: Valve area (Doppler): --- cm², Valve area (planimetry): --- cm², Mean transvalvular gradient: 4 mm Hg, Estimated pulmonary pressure: 38/9 mmHg using estimated CVP of 10 mm Hg. Diastolic function: DTI: Septal E: 1.8, E1: 0.04, E/E1: 43; Lateral E: 1.8, E1: 0.07, E/E1: 26; A Dur: 166 msec, Deceleration Time: 249 msec, IVRT: --- msec. RV Function: TAPSE 1.4 (Normal > 1.5 cm), S' --- (Normal > 13 cm/s).  **Measurements:** Sinus of Valsalva: --- cm, Index: --- cm/m² (Normal < 2.2 cm/m²), Aortic root: 2.7 cm, Index: 1.5 cm/m² (Normal < 2.2 cm/m²), Ascending Aorta: 3.3 cm, Index 1.8 cm/m² (Normal < 2.2 cm/m²), Aortic arch: 3.3 cm, Index: 1.8 cm/m² (Normal < 2.2 cm/m²), Aortic valve excursion: --- cm (Normal 1.6 - 2.6), Left atrium: 5.1 cm, Index 2.8 cm/m² (Normal < 2.1 cm/m²), Left atrial volume: 62 ml/m² (Normal < 29 ml/m²), LV septum diastolic thickness: 9 mm (6-9 mm), LV posterior wall diastolic thickness: 9 mm (6-9 mm), Left ventricular dimension: End diastolic size: 57 mm (39-53 mm), End systolic size: 41 mm, EDV: 162 ml (67-155 ml), EDVi: 90 ml/m² (35-75 ml/m²), ESV: 75 ml (22-58 ml), ESVi: 42 ml/m² (12-30 ml/m²), SV: 87 ml, SVi: 48 ml/m², Visual LVEF: 50%, Modified Simpson's LVEF: 55%, 54%. |
| **5** | Height: 62 inches Weight: 112 pounds Indication: xx-year-old woman with vascular calcifications noted on CT scanning of the chest. She has hypertension, family history of heart disease and history of cigarette smoking. Procedure: The patient exercised on a treadmill for a total of 6.7 minutes, reaching stage 3 of the Bruce protocol, achieving an estimated workload of 7 METs. The heart rate was 88 bpm at baseline, and increased to 150 bpm at peak exercise, representing 90% of age-predicted maximal heart rate. The blood pressure response was normal . Resting blood pressure was 118/76 mmHg, and peak/nadir blood pressure was 200/90 mmHg. The patient did not have chest pain/symptoms during the procedure. The electrocardiogram did not show ST-segment changes diagnostic for ischemia. There was high QRS voltage on baseline EKG and a nonspecific ST segment abnormality with exercise. No arrhythmias were observed. Echocardiographic imaging showed normal wall motion at rest. With exercise, there was a normal increase in contractility of all myocardial segments and a normal decrease in left ventricular cavity size. Image quality was excellent.  IMPRESSION: The patient underwent a submaximal exercise study demonstrating a fair exercise capacity. The study is negative for ischemia by symptoms, nondiagnostic for ischemia by EKG interpretation, and negative for ischemia by echocardiographic imaging. Overall, the study speaks against the presence of ischemic heart disease. |

**Table S3: Prompt Template for Finetuning Llama2-70b**

|  | **Prompt Component** |
| --- | --- |
| Prefix | Given below is a post-2016 TTE report with a dictionary consisting of key-value pairs for various data, along with a random example of a pre-2016 TTE report. Generate a new report TTE which consists of the information from post-2016 TTE report and the dictionary of key-value pairs, structured in the pre-2016 TTE report style. Do not include any information from the pre-2016 TTE report. Use it just for a reference of the style the report is written in. If any 'nan' values are encountered in the dictionary, do not include it in the new TTE report. |
| Instruction | Findings: Hyperdynamic systolic function. No regional wall motion abnormalities. Visually estimated ejection fraction is 65 % . Severe diastolic dysfunction, consistent with restrictive physiology. Mid cavitary obstruction identified with a peak pressure gradient of 13 mmHg. No significant cavitary or outflow obstruction identified.   Normal right ventricular size. Normal systolic function. Unable to provide a reliable estimate of right ventricular systolic pressure due to trivial nature of tricuspid regurgitant jet.   Systolic anterior motion of the mitral valve chordae. Normal appearing mitral valve leaflets. Trivial mitral regurgitation.   No significant pericardial effusion.  No prior studies for comparison.  Conclusion: "ProcedureType": "Limited Transthoracic Echocardiogram", "Weight(Kg)": 78.0, "Weight(Lb)": 171.96057, "NoPericardialEffusion": 0.0, "Indications": "GILEAD;", "EpicReasonforStudy": "LIBERTY HCM Screening Echo", "index": 3, "echo_EarliestProc_delta": "0 days", "echo_before_proc": 1, "outpatient_echo": 1, "right_reason_for_echo": 1, "complete_tte_echo": 0, "eligible_echo": 0}" |
| Suffix (Example of a variation of report from the pre-2016 dataset) | Transthoracic Echo Report BSA: 2.01 m^2 BP: 136/77 HR: 72 EKG Procedure: COMPLETE 2D, DOPPLER AND COLOR Agitated Saline: Yes Contrast: None 3D Imaging of: Left Ventricle Technical Quality: Good IMPRESSION Normal left and right ventricular size and systolic function. Estimated ejection fraction estimated by 3D volumetric analysis is 66 Normal diastolic function. Mild left atrial dilatation. Early right-to-left shunt by agitated saline (bubble) study, suggesting intracardiac shunt. Aneurysmal interatrial septum. No signficant valvular abnormalities. No prior studies for comparison. FINDINGS LEFT VENTRICLE: Normal left ventricular size. Normal wall thickness. Normal systolic function. No regional wall motion abnormalities. Estimated ejection fraction estimated by 3D volumetric analysis is 66 %. Normal diastolic function. RIGHT VENTRICLE: Normal right ventricular size. Normal systolic function. Estimated right ventricular systolic pressure is 30 mmHg. ATRIA: Mild left atrial dilatation. Normal right atrial size. Early right- to-left shunt by agitated saline (bubble) study, suggesting intracardiac shunt. Aneurysmal interatrial septum. MITRAL VALVE: Normal appearing mitral valve leaflets. No significant mitral regurgitation. AORTIC VALVE: Trileaflet aortic valve. No aortic stenosis. No significant aortic regurgitation. TRICUSPID VALVE: Structurally normal tricuspid valve leaflets. Mild tricuspid regurgitation. PULMONARY VALVE: Structurally normal pulmonic valve leaflets. No pulmonary regurgitation. AORTA: Normal visualized portions of the aorta. INFERIOR VENA CAVA: Normal inferior vena cava. PERICARDIUM: No significant pericardial effusion. MEASUREMENTS 2D ECHO LV Diastolic Diameter PLAX 5.3 cm (4.1 - 5.7) LV Systolic Diameter PLAX 3.4 cm (2.0 - 4.0 IVS Diastolic Thickness 0.7 cm (0.6 - 1.0 ) LVPW Diastolic Thickness 0.9 cm (0.6 - 1.0) LA Systolic Diameter LX 3.8 cm (3.0 - 4.0) LA Volume MOD BP Index 38.4 ml/m? (16 - 28 cm?/m?) Aortic Root Diameter 2.9 cm (2.7-4.1) DOPPLER LVOT Peak Velocity 1.3 m/s AV Peak Velocity 1.6 m/s (1.0-1.7 m/s) TR Peak Velocity 2.3 m/s TR Peak Gradient 21.4 mmHg (< 25 mmHg) Mitral E Point Velocity 1.1 m/s (0.6-1.3) Mitral A Point Velocity 0.7 m/s (0.3 -0.6) Mitral E to A Ratio 1.6 MV Deceleration Time 217.0 ms LV E' Lateral Velocity 12.2 cm/s (>=10cm/s) LV E' Septal Velocity 12.4 cm/s (>=8cm/s) Mitral E to LV E' Lateral Ratio 8.8 (=<8) Mitral E to LV E' Septal Ratio 8.6 (=<8) |
| Result | Limited Transthoracic Echocardiogram: Agitated Saline: Yes Contrast: Hyperdynamic systolic function. No regional wall motion abnormalities. Estimated ejection fraction 65%. Severe physiological diastolic function. Mild cavitary obstruction identified. Peak pressure gradient of 13 mmHg. No signficant cavitary outflow obstruction. FINDINGS RIGHT VENTRICLE: Normal right ventricular size. Normal systolic function. Tricuspid regurgitant jet. No reliable estimate of systolic pressure. MITRAL VALVE: Systolic anterior motion of mitral valve. Normal appearing mitral valve leaflets. Trivial mitral regurgitation. PERICARDIUM: No significant pericardial effusion. MEASUREMENTS 2D ECHO Ejection Fraction: 65%. Peak pressure gradient: 13 mmHg |

**Table S4: Prompt Template for Finetuning Llama2-13b**

|  | **Prompt Component** |
| --- | --- |
| Prefix | Convert the report given below to structured format for the columns 'GLS%','IVSd','LVDiastolicFunction','AVStructure','AVStenosis','AVRegurg','AIPHT','LVOTPkVel','LVOTPkGrad','MVStructure','MVStenosis','MVRegurgitation','EF','LVWallThickness', 'AVPkVel(m/s)', 'AVMnGrad(mmHg)', 'AVAContVTI', 'AVAIndex'. Give the result in json format with key-value pairs. If any value for a key is not found in the data, use 'nan' to fill it up. Donot fill up data that is not present in the given report. |
| Instruction | Limited Transthoracic Echocardiogram: Agitated Saline: Yes Contrast: Hyperdynamic systolic function. No regional wall motion abnormalities. Estimated ejection fraction 65%. Severe physiological diastolic function. Mild cavitary obstruction identified. Peak pressure gradient of 13 mmHg. No signficant cavitary outflow obstruction. FINDINGS RIGHT VENTRICLE: Normal right ventricular size. Normal systolic function. Tricuspid regurgitant jet. No reliable estimate of systolic pressure. MITRAL VALVE: Systolic anterior motion of mitral valve. Normal appearing mitral valve leaflets. Trivial mitral regurgitation. PERICARDIUM: No significant pericardial effusion. MEASUREMENTS 2D ECHO Ejection Fraction: 65%. Peak pressure gradient: 13 mmHg |
| Result | {"GLS%": NaN, "IVSd": NaN, "LVDiastolicFunction": Severe, "AVStructure": NaN, "AVStenosis": NaN, "AVRegurg": NaN, "AIPHT": NaN, "LVOTPkVel": NaN, "LVOTPkGrad": NaN, "MVStructure": Normal Appearing Leaflets, "MVStenosis": NaN, "MVRegurgitation": Trivial, "EF": 65%, "LVWallThickness": NaN, "AVPkVel(m/s)": NaN, "AVMnGrad(mmHg)": NaN, "AVAContVTI": NaN, "AVAIndex": NaN} |

**Table S5: Performance of zero shot Llama2-13b.**

| Prefix | Convert the report given below to structured format for the columns 'GLS%','IVSd','LVDiastolicFunction','AVStructure','AVStenosis','AVRegurg','AIPHT','LVOTPkVel','LVOTPkGrad','MVStructure','MVStenosis','MVRegurgitation','EF','LVWallThickness', 'AVPkVel(m/s)', 'AVMnGrad(mmHg)', 'AVAContVTI', 'AVAIndex'. Give the result in json format with key-value pairs. If any value for a key is not found in the data, use 'nan' to fill it up. Donot fill up data that is not present in the given report. |
| --- | --- |
| Instruction | Limited Transthoracic Echocardiogram: Agitated Saline: Yes Contrast: Hyperdynamic systolic function. No regional wall motion abnormalities. Estimated ejection fraction 65%. Severe physiological diastolic function. Mild cavitary obstruction identified. Peak pressure gradient of 13 mmHg. No signficant cavitary outflow obstruction. FINDINGS RIGHT VENTRICLE: Normal right ventricular size. Normal systolic function. Tricuspid regurgitant jet. No reliable estimate of systolic pressure. MITRAL VALVE: Systolic anterior motion of mitral valve. Normal appearing mitral valve leaflets. Trivial mitral regurgitation. PERICARDIUM: No significant pericardial effusion. MEASUREMENTS 2D ECHO Ejection Fraction: 65%. Peak pressure gradient: 13 mmHg |
| Response | echo_echo_earliest_proc_delta:0 days,echo_echo_before_proc:1,echo_echo_outpatient_echo:0,echo_echo_right_reason_for_echo:1,echo_echo_complete_tte_echo:1,echo_echo_eligible_echo:0,echo_echo_echo_procedure_date:,echo_echo_echo_procedure_time:,echo_echo_echo_procedure_duration:00:00:00,echo_echo_echo_procedure_status:Final,echo_echo_echo_date:,echo_echo_echo_procedure_type:Limited Transthoracic Echocardiogram,,echo_echo_echo_base_line_rhythm:,echo_echo_echo_reading_priority:,echo_echo_echo_ivsd:,echo_echo_echo_lvidss:,echo_echo_echo_lvidd,echo_echo_echo_lvpwd,echo_echo_echo_rvidd:,echo_echo_echo_laid2d,echo_echo_echo_lavol,echo_echo_echo_lavol_indexed,echo_echo_echo_mvpke:,echo_echo_echo_mvpka:,echo_echo_echo_e_medial |

**Table S6. Analytic packages and language used for model finetuning and evaluation.**

| **Programming Language/Package** | **Version** |
| --- | --- |
| Python | 3.12.1 |
| pandas | 2.2.1 |
| numpy | 1.26.4 |
| huggingface_hub | 0.20.3 |
| transformers | 4.38.1 |
| trl | 0.7.1 |
| peft | 0.5.0 |
| pytorch | 2.2.1 |
| scipy | 1.12.0 |
